# Hypoxia-Associated Gene Signature for Uterine Cervical Cancer

**DOI:** 10.64898/2026.03.20.26348602

**Authors:** Anubhav Datta, Luisa Vanesa Biolatti, Mark Reardon, Kamilla Bigos, Sapna Lunj, Helena Eke, Sudha Desai, Paula Hyder, Kimberley Reeves, Lisa Barraclough, Kate Haslett, Christina S Fjeldbo, Heidi Lyng, James P.B. O’Connor, Catharine M.L. West, Peter Hoskin, Ananya Choudhury

**Author notes:** Joint first authors (Anubhav Datta, Luisa Vanesa Biolatti, Peter Hoskin, Ananya Choudhury). Joint senior authors. **Correspondence**, Postal address : Dr Anubhav Datta, Clinical Radiology, The Christie NHS Foundation Trust, 550 Wilmslow Road, Manchester M20 4BX. United Kingdom.

## Abstract

**Background:** Tumour hypoxia is a major determinant of treatment resistance and poor prognosis in cervical cancer but remains difficult to assess in clinical practice. Gene expression signatures offer a potential means to characterise hypoxia-related biology. This study aimed to develop and validate a hypoxia-associated gene expression signature for cervical cancer.

**Methods:** RNA sequencing was performed on five cervical cancer cell lines exposed to normoxia (21% O₂) and hypoxia (1% O₂). Differentially expressed genes were mapped to The Cancer Genome Atlas cervical cancer cohort (TCGA-CESC) to train a 55-gene hypoxia classifier using k-means clustering and Prediction Analysis for Microarrays. The model was validated in an institutional Manchester cohort (n=153) and two public datasets from Seoul (n=300) and Oslo (n=283).

**Results:** The Manchester 55-gene signature was enriched for canonical hypoxia pathways. In the Manchester cohort, hypoxia classification correlated with advanced FIGO stage, nodal involvement, tumour size ≥ 4 cm, and hydronephrosis (*adjusted p*<0.05). Hypoxic tumours showed reduced overall survival (OS) and progression-free survival (PFS) in all cohorts. In multivariable models, the signature remained independently prognostic for OS in both TCGA (HR 1.70, 95% CI 1.10–2.60, *p*=0.012) and Manchester (HR 1.95, 95% CI 1.08–3.51, *p*=0.026). A direct comparison with a published 6-gene hypoxia signature in the Oslo cohort demonstrated 71% concordance in classification.

**Conclusions:** Our 55-gene signature should be tested prospectively in trials to assess its ability to stratify patients for hypoxia-targeted therapies.

## Introduction

Cervical cancer represents a major global health challenge, particularly in low- and middle-income countries where the burden is disproportionately high. In 2022, there were approximately 660,000 new cases and 350,000 deaths worldwide, making cervical cancer the fourth most common cancer among women ^1^. Despite the success of preventative strategies such as human papillomavirus (HPV) vaccination and screening programs, disparities in access to these interventions mean that many women present with advanced-stage disease requiring multimodality treatment ^2^.

The standard-of-care for locally advanced cervical cancer (LACC) involves definitive chemoradiotherapy, typically consisting of external beam radiotherapy (EBRT) combined with cisplatin-based chemotherapy, followed by intracavitary brachytherapy ^3^. Although treatment outcomes improved with modern radiotherapy techniques and image-guided brachytherapy, five-year survival rates remain suboptimal, particularly in patients with bulky or hypoxic tumours ^4,5^.

Tumour hypoxia – defined as oxygen deficiency within the tumour microenvironment (pO₂ ≤10 mmHg)^6^ – is a major adverse prognostic factor. Hypoxic tumours are associated with increased aggressiveness, metastasis, and resistance to radiotherapy and chemotherapy ^7^. Several therapeutic strategies have been developed to overcome hypoxia-related resistance in LACC. However, clinical trials assessing hypoxia-targeted therapies in cervical cancer yielded mixed results, often limited by the inability to accurately and reliably stratify patients based on the degree of tumour hypoxia ^8^. Thus, reliable assessment of hypoxia is critical for patient selection and therapeutic optimisation.

While multiple methods exist for assessing tumour hypoxia, each has practical limitations ^5^. Gene expression profiling is attractive, as hypoxia-associated gene signatures capture the transcriptional consequences of oxygen deprivation, integrating signals across many genes rather than relying on single markers. Several hypoxia gene signatures have already been proposed in cervical cancer, including an imaging-anchored model ^9^ and bioinformatics-based signatures derived from TCGA data ^10–12^. These studies highlight the promise of transcriptomic classifiers, but most have been validated in limited retrospective settings, leaving uncertainty about their robustness and generalisability across populations. Addressing this remains a key need for the field.

The aim of this study was to develop and validate a hypoxia-associated gene expression signature in patients with cervical cancer. Specifically, we sought to (i) identify a panel of genes responsive to experimentally defined hypoxia, (ii) construct a gene signature associated with poor prognosis, and (iii) validate the prognostic relevance of the signature in independent patient datasets.

## Materials and methods

### Cell line culture and hypoxia exposure

Cell line experiments were used to generate RNA for differential gene expression (DEG) analysis and to identify the candidate signature genes. Five cervical cancer cell lines (CaSki, HeLa, MS-751, SiHa and SW-756) were chosen to represent the disease as observed within the population. Cell lines used in the experiments were authenticated and mycoplasma tested by the Molecular Biology Core Facility, CRUK Manchester Institute, Alderley Park, UK. The cell lines were cultured according to established lab protocols ^13^ (Supplementary Material 1 – Table S1.1). In brief, cells were exposed to three different oxygen environments for 24 h – 21% O_2_ (normoxia) and 1% (Ruskin Invivo2 400 hypoxia workstation, Ruskin Technology Ltd, Bridgend, UK). Experiments were repeated for three different passages for each cell line.

### Cell line RNA extraction and RNA-seq analysis

RNA was extracted using the RNeasy Plus Mini Kit (Qiagen, Hilden, Germany) as per the manufacturer’s protocol. RNA quantity was assessed using the Qubit RNA broad range assay kit (Invitrogen, Thermo Fisher Scientific, Massachusetts, USA) to ensure a minimum concentration of 1 µg total RNA in 20 µl of RNase-free water. Extracted total RNA was processed by the Genomic Technologies Core Facility at the University of Manchester (Manchester, UK).

Libraries were prepared using the TruSeq Stranded mRNA assay (Illumina, San Diego, USA) according to the manufacturer’s protocol and sequenced using the Illumina HiSeq 4000 platform (Illumina, San Diego, USA), 76 base pair, paired end reads. Reads were aligned to the human reference genome (GRCh38.p13/hg38) ^14^ using STAR (v2.7.7a) ^15^. DEG analysis was performed on RNAseq data using DESeq2 (v 1.28.1) ^16^. Raw counts were normalised to the geometric mean across samples. Outlier genes were excluded in datasets with ≤6 replicates.

### Clinical cohorts

Data for 307 patients from The Cancer Genome Atlas Cervical Squamous Cell Carcinoma and Endocervical Adenocarcinoma (TCGA-CESC) cohort (upload date 28/01/2016) were downloaded via the Broad institute Firehose portal (https://gdac.broadinstitute.org/). Patients with locally advanced (stages 1B_2_ to IVA) squamous cell cervical cancer and whole transcriptome data (n=141) were selected. The mean age was 50±15 years (μ ± SD) and patients were treated with chemoradiation delivered with curative intent. This dataset was split into training (n=71) and validation (n=70) cohorts.

A retrospective patient cohort was curated at our institution as part of The ‘<u>Bio</u>markers for <u>C</u>linical <u>H</u>ypoxia <u>E</u>valuation in <u>C</u>ervical <u>C</u>ancer’ (BioCHECC) study (NCT05029258). The Manchester, U.K., cohort was used for external validation. Ethical approval was obtained (REC 20/NW/0377). Patients with carcinoma (squamous cell, adeno or adenosquamous) of the uterine cervix treated between 2013 and 2018 at The Christie NHS Foundation Trust were identified through the gynae-oncology database. Inclusion criteria were women >18 years with no upper age limit and biopsy-confirmed uterine cervix cancer. Women treated with curative intent (surgery or radical chemoradiation) were included. Surgical management involved radical hysterectomy ± bilateral salpingo-oophorectomy. Patients in the (chemo)radiotherapy sub-group received 25 fractions/5 weeks of external beam radiotherapy (EBRT) given to a total dose of 45 Gy (1.8 Gy per fraction). In keeping with protocol, clinical target volume (CTV) was defined to include gross tumour volume (GTV), entire cervix, entire uterus, parametria, ovaries and vagina depending on involvement^17^. Nodal CTV included the pelvic nodes. In addition, patients received two fractions of pulsed-dose rate brachytherapy delivered with an intracavitary or a combined intracavitary/interstitial implant. MRI-guided adaptive brachytherapy (IGABT) was prescribed to a dose of 40 to 45 Gy to reach a total chemoradiotherapy + IGABT dose of 85 to 90 Gy EQD_2_ to the high-risk CTV, and ≥60 Gy to the intermediate-risk CTV. Single-agent concomitant cisplatin chemotherapy was given weekly at 40 mg/m^2^.

Further external validation was performed in two publicly available cohorts. A patient cohort (GSE44001; Seoul, South Korea ^18^) was downloaded and processed in-house. The other patient cohort (GSE72723; Oslo, Norway ^9^) was used for independent validation as part of an inter-laboratory collaboration.

### Manchester cohort RNA extraction, and microarrays analysis

RNA was extracted from FFPE samples and used to validate the gene expression signature. Haematoxylin and eosin (H&E) stained sections of the FFPE tumour blocks were double reported by two consultant histopathologists (SD and PH) for histology, grade, and tumour surface area (TSA). The minimum required TSA for inclusion was 25%. RNA extraction was performed in collaboration with Histology Services, CRUK Manchester Institute, Alderley Park, UK, by use of the ‘Roche High Pure FFPE RNA Isolation Kit’ (06650775001; Basel, Switzerland). RNA quantity was assessed using the Qubit RNA broad range assay kit (Invitrogen, Thermo Fisher Scientific, Massachusetts, USA) to ensure a minimum concentration of 72 ng total RNA in 9 µl of RNase-free water. Quality was assessed using a 2200 TapeStation (Agilent Technologies, Santa Clara, USA) to ensure RNA integrity (RIN) values of at least 2.

Sample preparation and amplification followed the GeneChip 3’ IVT Pico Kit protocol. RNA was plated on a Clariom S plate (96-wells; 92 tumour samples and 4 controls) and processed by YourGene Health (Manchester, UK). Arrays were run on the GeneTitan Multi-Channel Instrument and hybridisation quality was checked using the Transcriptome Analysis Console (v 4.0.1, Applied Biosystems).

Raw image data (*.CEL files) were processed using the apt-probeset-summarize programme (v 1.20.0, Applied Biosystems) to yield log_2_ expression levels. Normalisation was performed within *apt-probeset-summarize.* Batch effects were corrected using the ComBat function in R/surrogate variable analysis (sva) ^19^. DEG were calculated using edgeR (v 2.26.0) ^20^.

### Candidate gene selection and signature threshold

Differential expression was defined as genes which displayed **|**≥2**|**-fold change between 21% and 1% oxygen. The TCGA data was used to define the hypoxia signature classification threshold. Candidate gene mapping to TCGA datasets was performed using Ensembl gene identifiers via the Ensembl genome database (release 100, GRCh38) through biomaRt R package (version 2.42.1; ^21^). Unsupervised *k*-means clustering (*k*=2) was used to partition a subset of the data (TCGA train) into two classes (Low and High Hypoxia) based on the candidate gene expression profiles. The labelled ‘TCGA train’ sub-group was used to train the Prediction Analysis for Micro-arrays (PAM) method ^22^. The PAM model was applied to the median-centred expression matrix to predict hypoxia status. Each sample was assigned to the nearest unshrunken centroid based on Spearman distance, yielding a binary classification of hypoxia low or hypoxia high. The final signature was tested in the TCGA test subset.

### Statistical analysis

All data analysis for gene expression related work was performed in R version 4.5.0 ^23^. False discovery rate correction was done using the Benjamini-Hochberg method ^24^. Study group proportions were compared using the Chi-squared tests and Fisher’s exact tests compared discrete variables.

Progression free survival (PFS) and overall survival (OS) were defined as time from diagnosis to last follow up or event occurrence (locoregional recurrence, metastasis, or death). All follow up data was censored at 5 years. Time to event analyses were performed using the Kaplan–Meier method and differences compared using the log-rank test.

Univariate and multivariate regression analyses for overall survival were conducted using hypoxia status as defined by the gene signature, age, performance status, histology, stage, differentiation, LVSI, size of tumour, parametrial invasion, lymph node involvement and hydronephrosis. Hazard ratios (HR) and 95% confidence intervals (CI) were reported in keeping with the Cox proportional hazard model. For categorical variables with sparse levels, categories with low frequencies were either collapsed into clinically or statistically appropriate groups or excluded altogether to ensure model stability and interpretability.

## Results

### Identification of the Manchester 55-Gene Hypoxia-Associated Signature

A total of 402 DEGs were identified between 21% and 1% oxygen (False Discovery Rate (FDR) corrected p < 0.1) in all 5 cell lines (Supplementary Material 1 – Table S1.1). A matrix discovery of FDR corrected p value (rows), and numbers of cell lines included (columns), was used to identify a 61 ‘candidate’ gene list (p<0.000001) based on differential expression in ≥4 cell lines (Supplementary Material 2 – Table S2.1). This approach has been used previously in hypoxia-related transcriptomic studies to balance stringency and coverage, requiring consistent regulation across most cell lines while allowing for some biological variability^25^. Of the 61 candidate genes, 55 were successfully mapped to the TCGA dataset (Supplementary Material 3 – Table S3.1). Six genes were not mapped due to differences in the human genome reference assemblies used to curate the two datasets (GRCh37 vs GRCh38).

All 55 candidate genes were up regulated under hypoxic conditions. The gene set includes established hypoxia-associated genes such as *CA9, VEGFA, BNIP3, EGLN1, LDHA*, and *HK2*, as well as additional genes implicated in cellular metabolism, transcriptional regulation, and stress response. Gene ontology (GO) enrichment analysis was performed to identify overrepresented biological processes within the gene set (Supplementary Material 3 – Figure S3.1). The most significantly enriched GO terms were “response to hypoxia,” “response to decreased oxygen levels,” and “response to oxygen levels,” all with adjusted p-values <1×10⁻⁷. Enrichment was also observed for metabolic processes including “glycolytic process,” “pyruvate metabolic process,” and “hexose metabolic process,” consistent with known hypoxia-driven metabolic reprogramming. Notably, the gene set showed strong enrichment for the “cellular response to hypoxia” term (fold enrichment = 45.24, p = 1.87 × 10 ^⁻09^, Supplementary Material 3 – Table S3.2), supporting the biological relevance of the selected genes.

### Hypoxia classification and internal validation

Using the expression profile of the 55 genes, unsupervised k-means clustering (*k*=2) was applied to the TCGA training cohort, resulting in two distinct groups (Figure 1a) with a clear differential gene expression. The high expression cluster (Class 2) was associated with the up-regulated candidate genes.

**Figure 1.**
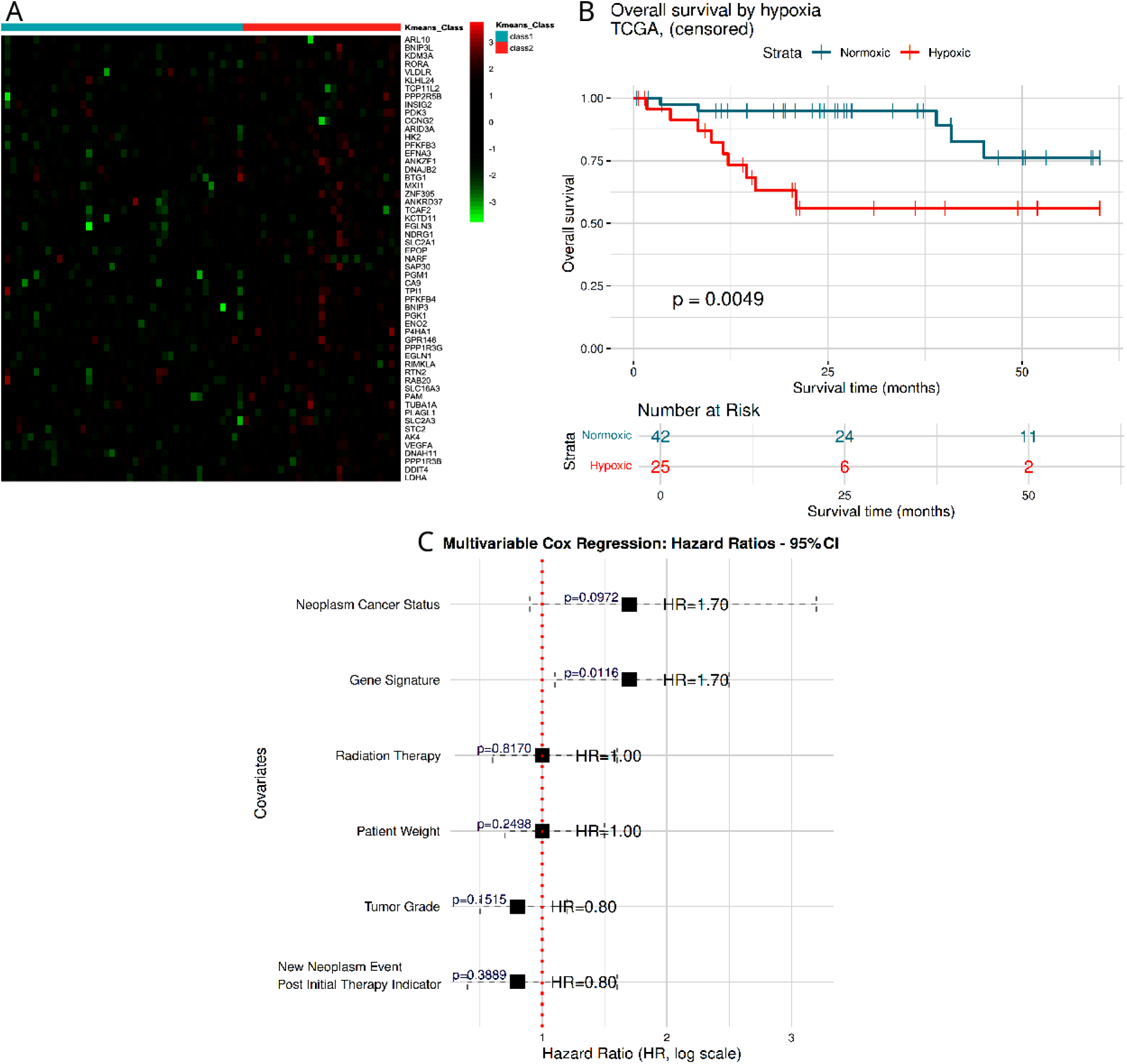
Application of the Manchester 55-gene signature to the TCGA cervical cancer dataset. (A) Heatmap displaying the expression of 55 hypoxia-associated genes in the TCGA training cohort (n = 70). Rows represent genes and columns represent tumour samples. Gene expression was scaled by row (z-score); red indicates high expression and green low expression. Unsupervised k-means clustering classified tumours into two groups: Class 1 (pale blue) and Class 2 (pale red). (B) Kaplan-Meier analysis of overall survival in the TCGA internal validation subset (n = 67). Twenty-five tumours were classified as hypoxic by the gene signature, and these demonstrated significantly worse overall survival compared to normoxic tumours (log-rank p = 0.0049). (C) Multivariable Cox proportional hazards model assessing the association between hypoxia status and overall survival in the full TCGA cohort, adjusting for clinical covariates. Hypoxia classification remained an independent predictor of poorer survival (HR 1.7, p = 0.012). Horizontal bars indicate 95% confidence intervals; the vertical dashed line represents the null (HR = 1).

The TCGA training set was used to derive the PAM classifier. The optimal model size, selected via 10-fold cross-validation, minimised classification errors and was significantly associated with a poor prognosis in the TCGA cohort. The Manchester 55-gene signature achieved a cross-validated misclassification error of <5% and 93% accuracy in independent cell line validation. The model was tested on the TCGA test set, where it stratified patients into hypoxic and normoxic groups. Kaplan-Meier (Figure 1b) show patient samples classified as hypoxic had worse OS (p=0.0049) and PFS (p=0.0063). In a multivariable Cox proportional hazards model adjusting for tumour grade, radiation therapy, recurrence, weight, and neoplasm status (new neoplasm event that represents recurrence), the hypoxia classification remained an independent prognostic factor for poor survival (HR 1.7, p = 0.012; Figure 1C). Additional results of back testing in the cell line data and co-expression analysis in the TCGA data are presented in Supplementary Material 4 – Figure S4.1, Figure S4.2, Figure S4.3.

### External validation in the Manchester cohort

A total of 373 patients were identified and 153 biopsy samples were processed. Supplementary Material 5 – Figure S5.1, Figure S5.2, Figure S5.3). Table 1 summarises the clinicopathologic characteristics of the included 153 patients. Tumour hypoxia was associated with advanced clinical stage (adjusted *p* = 0.0010), larger tumour size (≥4 cm; adjusted *p* = 0.0016), pelvic lymph node involvement (adjusted *p* = 0.0016), and the presence of hydronephrosis (adjusted *p* = 0.0321).

**Table 1.** Baseline characteristics of women in the Manchester cohort (n = 153). Study group proportions were compared using the chi-squared test, or Fisher’s exact test (simulated) when expected cell counts were <5. P values were adjusted for multiple comparisons using the Benjamini–Hochberg method.

| Parameter | Category | Negative | Positive | Test chosen | FDR (BH) |
| --- | --- | --- | --- | --- | --- |
| **Age** | <40 years | 29 | 12 | Chi-squared | 0.37<sup>ns</sup> |
|  | ≥40 years | 67 | 45 |  |  |
| **PS<sup>^</sup>** | ≥2 | 6 | 5 | Fisher (simulated) | 0.35<sup>ns</sup> |
|  | 0 | 65 | 31 |  |  |
|  | 1 | 25 | 21 |  |  |
| **Clinical stage** | I | 38 | 4 | Fisher (simulated) | 0.001<sup>***</sup> |
|  | II | 49 | 34 |  |  |
|  | III | 4 | 9 |  |  |
|  | IV | 5 | 10 |  |  |
| **Histology** | Adeno | 30 | 9 | Chi-squared | 0.11<sup>ns</sup> |
|  | Squamous | 66 | 48 |  |  |
| **Grade** | ≥3 | 44 | 31 | Chi-squared | 0.55<sup>ns</sup> |
|  | 1 | 9 | 6 |  |  |
|  | 2 | 43 | 20 |  |  |
| **LVSI<sup>†</sup>** | Absent | 42 | 22 | Chi-squared | 0.14<sup>ns</sup> |
|  | Not known | 28 | 26 |  |  |
| **Tumour size** | Present | 26 | 9 | Chi-squared | 0.002<sup>**</sup> |
|  | <4cm | 39 | 7 |  |  |
|  | ≥4cm | 57 | 50 |  |  |
| **Pelvic nodes** | no | 63 | 20 | Chi-squared | 0.002<sup>**</sup> |
|  | yes | 33 | 37 |  |  |
| **Para-aortic nodes** | no | 92 | 55 | Fisher (simulated) | 1<sup>ns</sup> |
| **Hydro-nephrosis** | yes | 4 | 2 | Chi-squared | 0.03<sup>*</sup> |
|  | no | 90 | 45 |  |  |
|  | yes | 6 | 12 |  |  |

<sup>^</sup>PS = performance status; <sup>†</sup>LVSI = lymphovascular space invasion; ns=not significant

Kaplan-Meier survival analysis of the Manchester cohort showed that tumour hypoxia was associated with poorer clinical outcomes (Figure 2a and b). Median progression-free (PFS) and overall survival (OS) were not reached in either the normoxic or hypoxic groups. In the PFS analysis, patients with hypoxic tumours exhibited a significantly reduced probability of remaining progression-free compared to those with normoxic tumours (p = 0.042). Similarly, OS was significantly worse in the hypoxic group (p = 0.017). Five-year censored event rates for all patients are presented in Supplementary Material 6 – Table S6.1.

**Figure 2.**
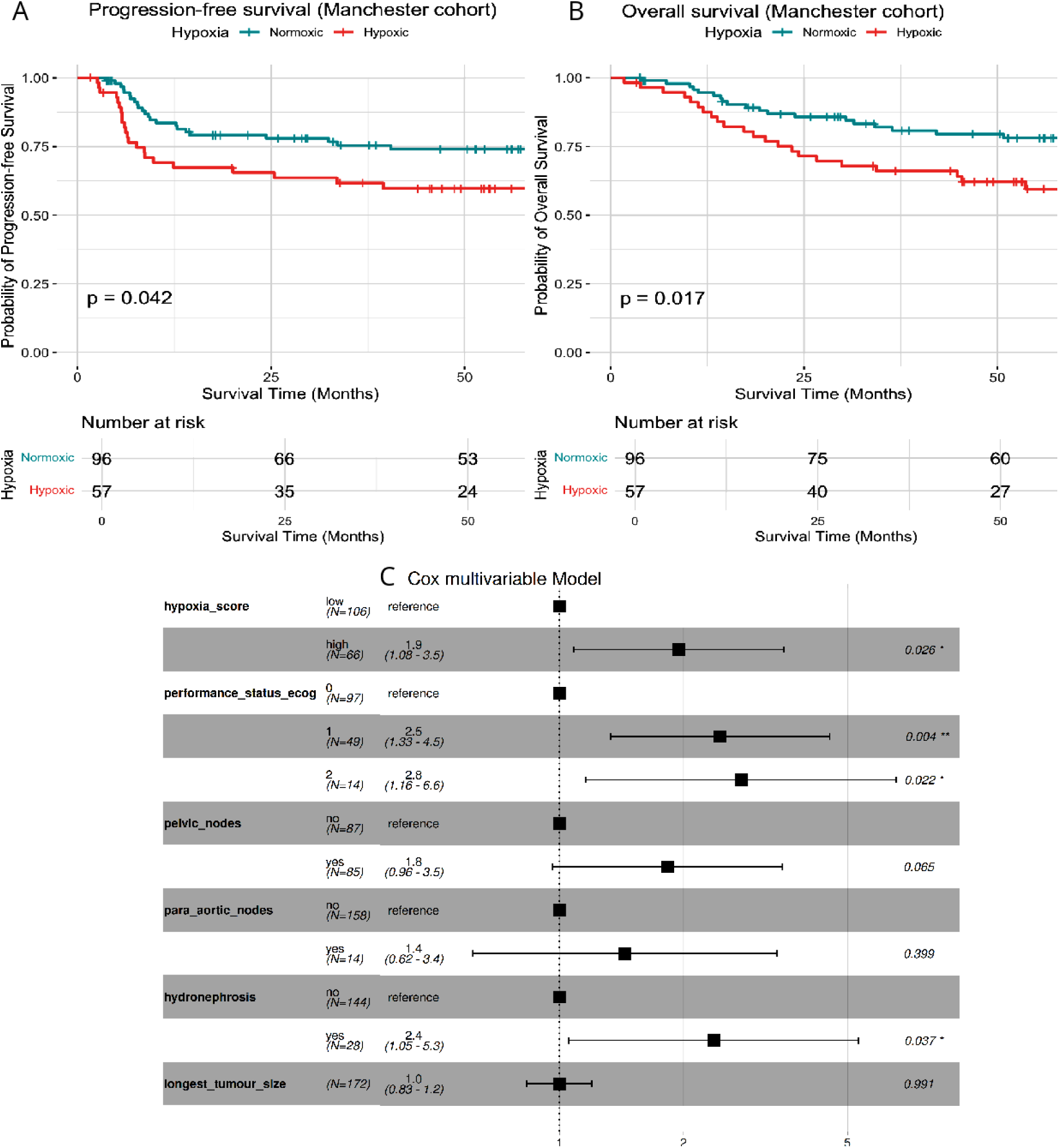
Kaplan-Meier survival curves and multivariate Cox regression analysis for overall survival. (A) Progression-free survival (PFS) and (B) overall survival (OS) stratified by tumour hypoxia status in the Manchester cohort. Patients with hypoxic tumours had significantly worse outcomes compared to normoxic tumours (PFS: p = 0.042; OS: p = 0.017). Tick marks indicate censored cases. (C) Forest plot of the multivariate Cox proportional hazards model for OS. Hazard ratios (HRs) and 95% confidence intervals (CIs) are shown for covariates included in the model. A vertical dashed line indicates the null value (HR = 1.0).

On univariate analysis (Table 2), a high hypoxia score associated with inferior OS (HR 2.72, 95% CI 1.68–4.40, p = 4.6 × 10⁻⁵), alongside poor performance status, nodal involvement, hydronephrosis, and larger tumour size. In the multivariable model (Figure 2c), high hypoxia score remained an independent prognostic factor (HR 1.95, 95% CI 1.08–3.51, p = 0.026), together with ECOG performance status (ECOG 1: HR 2.46, 95% CI 1.33–4.53; ECOG 2: HR 2.76, 95% CI 1.16–6.58) and hydronephrosis (HR 2.37, 95% CI 1.05–5.32, p = 0.037).

**Table 2.** Univariable Cox regression analysis of prognostic factors for overall survival.

| **Covariate** | **Level** | **HR** | **95% CI** | **P Value** |
| --- | --- | --- | --- | --- |
| **Hypoxia score** | High | 2.72 | 1.68 – 4.40 | 4.61e-05 |
| **Performance status (ECOG)** | 1 | 3.19 | 1.78 – 5.70 | 9.16e-05 |
| **Performance status (ECOG)** | 2 | 5.83 | 2.79 – 12.21 | 2.87e-06 |
| **Pelvic nodes** | Yes | 2.95 | 1.77 – 4.91 | 3.10e-05 |
| **Para-aortic nodes** | Yes | 4.46 | 2.36 – 8.40 | 3.82e-06 |
| **Hydronephrosis** | Yes | 5.37 | 3.26 – 8.84 | 3.89e-11 |
| **Longest tumour size** | — | 1.40 | 1.25 – 1.57 | 4.83e-09 |

### External validation in publicly available cohorts

Supplementary Material 7 – Table S7.1 summarises the clinical characteristics across the three cohorts. Figure 3 shows Kaplan–Meier survival analyses stratified by the 55-gene hypoxia signature across two independent cervical cancer cohorts. In the South Korean cohort (n = 300), patients classified as hypoxic (n = 59) had significantly reduced disease-free survival compared to normoxic patients (n = 241; p = 0.0025; Figure 3A). In the Norwegian cohort (n = 283), hypoxic patients (n = 109) also demonstrated significantly poorer outcomes. Progression-free survival was shorter in the hypoxic group compared to the normoxic group (n = 174; p = 0.0062; Figure 3B), and overall survival was similarly reduced in the hypoxic group (p = 0.0033; Figure 3C).

**Figure 3.**
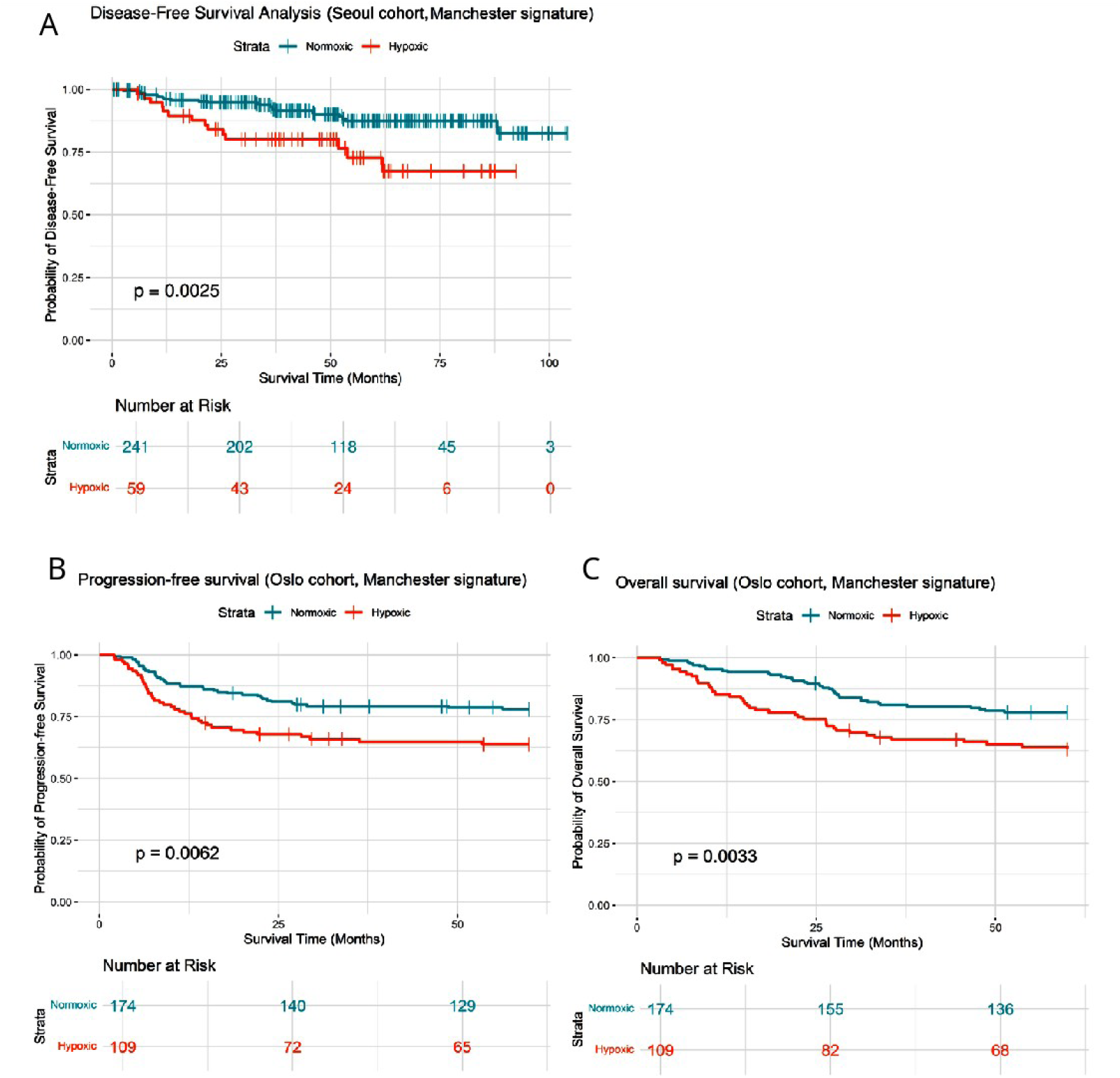
Kaplan-Meier survival curves stratified by hypoxia status, as defined by the Manchester gene signature, in two independent cervical cancer cohorts. (A) Disease-free survival in the South Korean cohort (GSE44001), showing significantly reduced survival among hypoxic patients (p = 0.0025). (B) Progression-free survival and (C) overall survival in the Norwegian cohort (GSE72723) demonstrate consistent findings, with significantly poorer outcomes in hypoxic tumours (p = 0.0062 and p = 0.0033, respectively). Shaded areas represent 95% confidence intervals, and tick marks denote censored observations.

Figure 4 compares hypoxia classification outcomes between the 55-gene Manchester signature and an alternative 6-gene Oslo signature ^9^ in the Norwegian cohort (n = 283). Concordant classifications were observed in 202 patients (71%), with 138 patients classified as less hypoxic and 64 as more hypoxic by both signatures. Discordant classifications were observed in the remaining 81 patients, including 45 identified as more hypoxic only by the Manchester signature and 36 identified as more hypoxic only by the Oslo signature.

**Figure 4.**
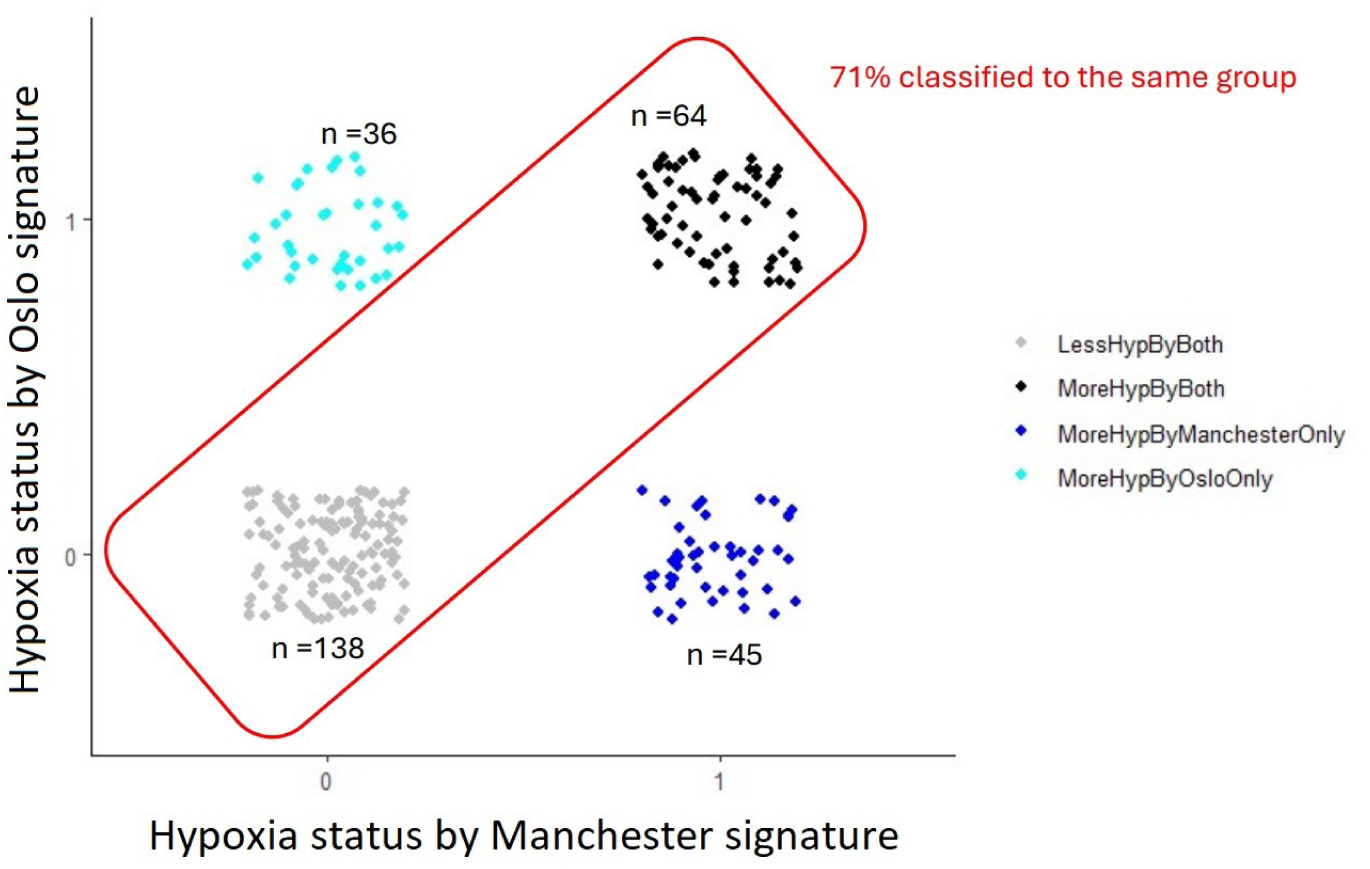
Comparison of hypoxia classification outcomes. Stacked bar chart displaying patient distribution by agreement status: ‘Less hypoxic’ (n=138) and ‘More hypoxic’ (n=64) by both signatures, versus discordant classifications (54-gene unique: n=45; 6-gene unique: n=36). Total concordance: 71% (n=202/283).

## Discussion

The primary findings of this study are threefold: (1) the de novo 55-gene expression signature showed significant enrichment for known hypoxia-related pathways; (2) the signature was associated with clinicopathological features of locally advanced and biologically aggressive cervical cancer; and (3) hypoxic classification by the signature was significantly associated with adverse survival across three independent patient datasets. Collectively, these findings support the potential of this signature to classify women with uterine cervical cancer according to clinical hypoxia status.

The gene set showed strong enrichment for canonical hypoxia pathways, particularly HIF-1α signalling and metabolic adaptation. It incorporated established hypoxia-responsive genes such as *VEGFA, CA9, SLC2A1/GLUT1*, and *LDHA*, together with AMPK-regulated metabolic genes (*PFKFB3, PFKFB4*) and stress-response mediators (*BNIP3, BNIP3L, NDRG1, PGK1*). These components collectively reflect complementary mechanisms of cellular adaptation to oxygen deprivation, spanning angiogenesis ^26^, glycolytic reprogramming ^27^, and autophagy ^28^. The fact that the genes cluster within well-established hypoxia pathways supports the idea that the signature reflects true tumour hypoxia rather than random variation in gene expression.

Comparison with 3 other published cervical cancer signatures: Fjeldbo ^9^ (6-gene), Yang ^10^ (5-gene) and Nie ^11^ (9-gene), show some similarities. Common genes across at least two signatures included *AK4*, *DDIT3/DDIT4*, *HK2*, *LDHA*, *P4HA1/P4HA2*, *PGM1*, *STC1/STC2*, and *VEGFA*, suggesting conserved hypoxia-related pathways across independent studies. The similarities suggest a conserved hypoxia-related pathway in cervical cancer (e.g. procollagen prolyl 4-hydroxylase domain ^29^) that maybe a target for treatment ^30^.

Differences in the signatures likely reflect varying model inputs and methodological approaches. The Fjeldbo classifier was derived by correlating gene expression with a magnetic resonance imaging parameter (ABrix). The Yang and Nie models were generated *in silico*, entirely within TCGA-CESC, using predefined hypoxia gene lists from MSigDB. Our approach began with *in vitro* oxygen modulation at 1% O₂, representing physiologically relevant hypoxia levels associated with radioresistance in cervical tumours ^6,31^, and only applied TCGA data for training and threshold determination. These differences highlight the lack of standardisation in hypoxia modelling and the importance of grounding classifier development in biological data.

Earlier efforts sought a single pan-cancer hypoxia metagene ^32,33^. However, most gene signatures have since proved single cancer type specific ^34–36^ or even histological subtype specific ^25,37^. The Fjeldbo classifier was trained exclusively in squamous cell carcinomas (SCC), whereas the Yang and Nie signatures incorporated all histological subtypes within TCGACESC. In our cohorts, SCC accounted for roughly three-quarters of cases, reflecting the global incidence pattern ^38^. Although adenocarcinoma numbers were insufficient for robust subgroup analysis, the signature’s consistent prognostic performance across cohorts suggests generalisation beyond a single histology. Larger datasets will be required to test subtype-specific behaviour formally.

In the institutional Manchester cohort, hypoxia classification associated strongly with established markers of poor prognosis, including hydronephrosis, nodal involvement, and tumour size ^39^. On multivariable analysis, the signature retained independent prognostic value alongside clinical covariates, aligning with previous evidence that hypoxia contributes to radioresistance, metastasis, and treatment failure in cervical cancer ^40^. These findings reinforce the validity of the signature as an indicator of aggressive disease.

A unique strength of this study is the direct, patient-level comparison between two independently derived cervical cancer hypoxia signatures within the same cohort. To our knowledge, this represents the first head-to-head evaluation of transcriptomic hypoxia classifiers applied to identical patient samples. When both the 55-gene Manchester and 6-gene Oslo signatures were applied to the Norwegian cohort, concordant hypoxia classification was observed in 71 % of tumours (Figure 4). This degree of agreement indicates partial biological convergence, while discordant cases likely reflect differences in pathway breadth and discovery approach. The broader 55-gene model may capture a wider range of hypoxia-responsive processes, whereas the compact 6-gene panel may emphasise a narrower response profile typical of squamous carcinomas. This direct comparison provides new insight into the generalisability of hypoxia-associated gene signatures in cervical cancer.

Methodological considerations should be acknowledged. The absence of publicly available datasets containing direct hypoxia measurements (such as pimonidazole staining or pO₂ histography) required surrogate labelling using unsupervised clustering in TCGA. Although consistent with prior studies ^9–11^, this approach relies on inferred rather than measured hypoxia and therefore cannot be directly compared with established reference methods. Training the model on *in vitro* data ensured biological specificity but may not fully capture the temporal and spatial heterogeneity of hypoxia *in vivo*. Minor platform-related differences should also be recognised. One gene (*EPOP*) from the 55-gene panel was unavailable in the Oslo dataset, and expression data across cohorts originated from distinct array platforms and preprocessing pipelines. Consequently, absolute risk scores and p-values cannot be directly compared, and a pooled multivariable analysis was not performed. Nevertheless, the consistent direction and magnitude of prognostic associations across independent datasets support the robustness of the classifier across technically and geographically diverse cohorts. The retrospective nature of this study remains a limitation, and prospective validation in well annotated cohorts, ideally including hypoxia imaging or tissue markers, will be essential before clinical use.

The strengths of this work include its integration of experimental, institutional, and publicly available data, multi-cohort validation, and direct cross-signature assessment. These address key concerns about reproducibility that have limited translation of earlier hypoxia biomarkers ^32^. Clinically, the 55 gene signature could provide a practical way to stratify patients by tumour hypoxia using pre-treatment fixed formalin paraffin embedded biopsy material. This form of molecular classification may support risk adapted management, identifying patients who could benefit from hypoxia modifying therapy, treatment intensification, or image guided dose escalation. As molecular profiling becomes more common in clinical oncology, transcriptomic hypoxia assessment could complement immune or metabolic biomarkers to refine risk prediction and guide personalised treatment strategies.

## Conclusion

This study provides a framework for assessing the transcriptional response to tumour hypoxia in cervical cancer and demonstrates its reproducibility across independent datasets. The head-to-head comparison of two classifiers highlights the feasibility of benchmarking hypoxia gene expression signatures within the same cohort. Prospective validation in trials will be essential to determine whether these classifiers can help guide hypoxia targeted or intensified therapies.

## Additional information

## Acknowledgements

The research was carried out at the National Institute for Health and Care Research (NIHR) Manchester Biomedical Research Centre (BRC) (NIHR203308).

## Authors’ contributions

Each of the contributing authors meet all of the following criteria:

- Conceived and/or designed the work that led to the submission, acquired data, and/or played an important role in interpreting the results.
- Drafted or revised the manuscript.
- Approved the final version.
- Agreed to be accountable for all aspects of the work in ensuring that questions related to the accuracy or integrity of any part of the work are appropriately investigated and resolved.

In addition, the following contributions apply at an individual level:

- Methodology: AD, MR, SL, CW, PH, AC
- Formal analysis: AD, LB, MR, CF
- Data curation: AD, LB, MR, KB, SL, HE, SD, PH, LB, KH, PH
- Visualisation: AD, LB
- Supervision: PH, AC, CW, JPBOC, LB, SL, MR
- Funding acquisition: PH, AC, CW, JPBOC

## Ethics approval and consent to participate

The studies involving humans were approved by Northwest-Preston Research Ethics Committee: 20/NW/0377. The studies were conducted in accordance with the local legislation and institutional requirements. The participants provided their written informed consent to participate in this study. The study was conducted in accordance with the Declaration of Helsinki.

## Data availability

Study data is available at request.

